# Health impact of monoclonal gammopathy of undetermined significance (MGUS) and monoclonal B-cell lymphocytosis (MBL): findings from a UK population-based cohort

**DOI:** 10.1101/2020.05.30.20117549

**Authors:** Maxine Lamb, Alexandra G Smith, Daniel Painter, Eleanor Kane, Tim Bagguley, Robert Newton, Debra Howell, Gordon Cook, Ruth de Tute, Andrew Rawstron, Russell Patmore, Eve Roman

## Abstract

**Objective:** To examine co-morbidity patterns in individuals with monoclonal gammopathy of undetermined significance (MGUS) and monoclonal B-cell lymphocytosis (MBL), both before and after premalignancy diagnosis; and compare their activity to that of the general population.

**Design:** Population-based patient cohort, within which each patient is matched at diagnosis to 10 age and sex-matched individuals from the general population. Both cohorts are linked to nationwide information on deaths, cancer registrations, and Hospital Episode Statistics (HES).

**Setting:** The UK’s Haematological Malignancy Research Network; which has a catchment population of around 4 million served by 14 hospitals and a central diagnostic laboratory.

**Participants:** All patients newly diagnosed 2009–15 with MGUS (n = 2203) or MBL (n = 561), and their age and sex-matched comparators (n = 27,638).

**Main Outcome measures:** Survival, and hospital inpatient and outpatient activity in the five years before, and three years after, diagnosis.

**Results:** Individuals with MGUS experienced excess morbidity in the 5-years before diagnosis, and excess mortality and morbidity in the 3-years after. Increased rate-ratios (RR) were evident for nearly all clinical specialties; the largest, both before and after diagnosis, being for nephrology (before RR = 4.38, 95% Confidence Interval 3.99–4.81; after RR = 14.7, 95% CI 13.5–15.9) and rheumatology (before RR = 3.38, 95% CI 3.16–3.61; after RR = 5.45, 95% CI 5.09–5.83). Strong effects were also evident for endocrinology, neurology, dermatology and respiratory medicine. Conversely, only marginal increases in mortality and morbidity were evident for MBL.

**Conclusions:** From a haematological malignancy perspective, MGUS and MBL are generally considered to be relatively benign. Nonetheless, monoclonal gammopathy has the potential to cause systemic disease and wide-ranging damage to most organs and tissues. Hence, even though most people with monoclonal immunoglobulins never develop a B-cell malignancy or suffer from any other form of M-protein related organ/tissue related disorder, the consequences for those that do can be extremely serious.

## BACKGROUND

Monoclonal gammopathy of undetermined significance (MGUS) and monoclonal B-cell lymphocytosis (MBL) are premalignant monoclonal B-cell disorders; the former progressing to myeloma at a rate of around 1% per year[1,2] and the latter to chronic lymphocytic leukaemia (CLL) at around 2% per year.[3,4] Diagnosed more frequently in men than women and people over 60 years of age,[5,6] overt symptoms of haematological malignancy are, by definition, absent in both MBL and MGUS.[7] Accordingly, although some premalignant disorders are found coincidentally during routine health checks, others are identified during diagnostic work-up investigations; MGUS during the course of tests applied to detect a range of potential conditions and illnesses,[8,9] and MBL during episodes of unexplained lymphocytosis.[4,10]

In addition to the association with haematological malignancy, individuals with MGUS or MBL can experience higher than expected levels of mortality and morbidity that are independent of cancer.[4,8,11–16] Indeed, although most individuals with these disorders suffer no obvious ill effects, interest in their relationship with other co-morbidities has increased markedly in recent years; MBL largely in relation to its potential to impact on the immune response,[17] and MGUS due to the systemic organ and tissue damage that can be caused by monoclonal immunoglobins secreted by the abnormal B-cell clone.[18] Hitherto, however, most information about these associations has been derived either from case-control studies established to look at risk factors for disease development (e.g. family history of disease), additional tests applied to specific patient groups (e.g. patients with kidney disease), or cohort studies that track individuals with either MGUS/MBL forwards in time from their diagnosis.[5,13,18] However, despite the undoubted interest in the sequence of health events, as far as we are aware no systematic population-based investigations of the co-morbidity patterns that precede and succeed a diagnosis of either MGUS or MBL have been undertaken in the same cohort.

With a view to shedding light on the health events occurring before and after the diagnosis of MGUS and MBL, the present report uses data from an established UK population-based patient cohort of haematological malignancies and related disorders to examine the co-morbidity patterns of individuals with these premalignancy clonal disorders (MGUS = 2203, MBL = 561). To enable effect size quantification, these patterns are compared to the baseline activity of an individually age- and sex-matched (10 per patient) general population comparison cohort.

## METHODS

Cases are from the Haematological Malignancy Research Network (HMRN; www.hmrn.org), a specialist UK registry established in 2004 to provide robust generalizable data to inform contemporary clinical practice and research across the country as a whole.[19,20] Set within a catchment population of around four million that is served by 14 hospitals and has a socioeconomic profile which is broadly representative of the UK as a whole, all haematological cancers and related conditions are diagnosed and coded by clinical specialists at a single integrated haematopathology laboratory, the Haematological Malignancy Diagnostic Service www.hmds.info) using standardized diagnostic criteria and the latest WHO ICD-O-3 classification.[7] Specifically, in relation to the present report, MBL was defined by a peripheral blood monoclonal B-cell count <5 ×10^9^/L and MGUS by the detection of paraprotein in peripheral blood and/or neoplastic lymphoplasmacytic/plasma cells in the bone marrow.

To facilitate comparisons with the general population, HMRN also has a general population cohort; all patients diagnosed 2009–15 were individually matched on age and sex to 10 unaffected individuals from the same catchment population, each one of which was assigned a pseudo-diagnosis date corresponding to the date of diagnosis of their matched case.[21,22] HMRN operates under a legal basis that permits data to be collected directly from clinical records without explicit consent; and all individuals in the patient cohort and the comparison cohort are linked to nationwide information on deaths, cancer registrations and Hospital Episode Statistics (HES).

Using similar methods to those previously described,[21–23] associations with hospital inpatient activity (HES Admitted Patient Care; HES-APC) and outpatient activity (HES Outpatient; HES-OP) in the five years prior to diagnosis/pseudo-diagnosis through to the three years after diagnosis were investigated. HES inpatient data contains ICD-10 (International Statistical Classification of Diseases 10^th^ revision) illness codes derived from discharge summaries[24]; and associations with these were examined in relation to the 17 specific conditions in the Charlson Comorbidity Index.[25–27] By contrast, HES outpatient data contains details about the type of outpatient attendance; the majority being linked to consultant specialty codes (e.g. ophthalmology, rheumatology etc.), with the remainder largely comprising routine follow-up/monitoring, nurse-led clinic attendances (e.g. anticoagulant clinics) and consultations with allied health professionals (e.g. podiatry).

This report includes all patients (cases) who were newly diagnosed with either MGUS (n = 2203) or MBL (n = 561) between 1^st^ January 2009 and 31^st^ December 2015 and their matched controls (n = 27,638); those diagnosed with a haematological cancer within six months of their MGUS/MBL diagnosis were considered ineligible. All cases and controls were followed-up for cancer registration and death until March 2017, and hospital activity (inpatient and outpatient) until March 2016. Additionally, progressions and/or transformations in cases were identified through HMDS up to March 2017. Data were summarised using standard methods. Overall survival, hospital activity and rate ratios (RRs) were calculated using time-to-event analyses. The Stata programme ‘strel’ was used to estimate relative survival (RS), using age- and sex-specific background mortality rates from national life tables.[28,29] All analyses were conducted using Stata 16.0.

### Patient and public involvement (PPI)

PPI is integral to HMRN, and takes place via a dedicated partnership, overseen by a lay committee. Patients from the partnership are involved in identifying key research questions and participate in all our funding applications. Furthermore, patients and their relatives routinely take part in the dissemination of HMRN’s findings, which also occurs via our lay website: www.yhhn.org

## RESULTS

Characteristics of individuals with either MGUS or MBL are summarized alongside their corresponding controls in Table 1. Both MGUS and MBL were more commonly diagnosed in males (51.6% MGUS, 58.5% MBL) and individuals over the age of 70 years (median diagnostic age 73.3 years for MGUS and 72.1 years for MBL). At 90.0% (95% Confidence Interval 87.2–92.3%), the 5-year relative survival (RS) of patients diagnosed with MGUS was significantly lower (p<0.01) than that of their general population controls (RS 99.5%, 95% CI 98.7–99.8%). For MBL, the difference was far less marked; the 5-year RS being 94.3% (95% CI 86.3–97.7%) for MBL cases and 98.5% (95% CI 96.8–99.3%) for their controls. Interestingly, the observed variations in blood cancer progression were in the opposite direction to those seen for mortality: 75 (3.4%) MGUS patients were diagnosed with a haematological malignancy (48/75 were myelomas) before April 2017, compared to 140 (25.0%) MBL patients (137/140 were CLLs). No significant differences were, however, evident for non-haematological cancer registrations; although the frequencies were slightly higher for MGUS (8.4%) and MBL (10.3%) than for their corresponding controls (8.0% and 8.8% respectively).

**Table 1:** Characteristics of patients diagnosed with monoclonal gammopathy of undetermined significance (MGUS) or monoclonal B-cell lymphocytosis (MBL) and their corresponding controls; HMRN diagnoses 2009–2015

|  |  | MGUS |  | MBL |  |
| --- | --- | --- | --- | --- | --- |
|  |  | Cases<br>Number (%) | Controls<br>Number (%) | Cases<br>Number (%) | Controls<br>Number (%) |
| **Total** |  | 2203 (100) | 22028 (100) | 561 (100) | 5610 (100) |
| **Gender** | **Male** | 1136 (51.6) | 11358 (51.6) | 328 (58.5) | 3280 (58.5) |
|  | **Female** | 1067 (48.4) | 10670 (48.4) | 233 (41.5) | 2330 (41.5) |
| **Age (years)** | **<60** | 349 (15.8) | 3490 (15.8) | 85 (15.2) | 850 (15.2) |
|  | **60-70** | 502 (22.8) | 5020 (22.8) | 153 (27.3) | 1530 (27.3) |
|  | **70-80** | 788 (35.8) | 7880 (35.8) | 194 (34.6) | 1940 (34.6) |
|  | **≥80** | 564 (25.6) | 5638 (25.6) | 129 (23.0) | 1290 (23.0) |
|  | **Median (IQR)** | 73.3 (64.6 - 80.2) | 73.3 (64.6 - 80.2) | 72.1 (64.5 - 79.2) | 72.1 (64.5 - 79.2) |
| **Overall survival<sup>1</sup>** | **5-year (95% CI)** | 71.8 (69.5-74.0) | 80.1 (79.5-80.8) | 77.2 (72.5-81.2) | 80.7 (79.4-81.9) |
| **Relative survival** | **5-year (95% CI)** | 90.0 (87.2-92.3) | 99.5 (98.7-99.8) | 94.3 (86.3, 97.7) | 98.5 (96.8-99.3) |
| **Progressions and Transformations<sup>1</sup>** | **Total** | 75 (3.4) |  | 140 (25.0) |  |
|  | **Myeloma** | 48 (2.2) | - | - | - |
|  | **CLL<sup>2</sup>** | 1 (0.05) | - | 137 (24.4) | - |
|  | **Other** | 26 (1.2) | - | 3 (0.5) | - |
| **Non-haematological cancer registrations<sup>1</sup>** | **Total** | 186 (8.4) | 1759 (8.0) | 58 (10.3) | 494 (8.8) |
|  | **Lung** | 32 (1.5) | 191 (0.9) | 4 (0.7) | 59 (1.1) |
|  | **Prostate<sup>3</sup>** | 20 (1.8) | 179 (1.6) | 3 (0.9) | 56 (1.7) |
|  | **Colorectal** | 19 (0.9) | 167 (0.8) | 6 (1.1) | 40 (0.7) |
|  | **Breast<sup>3</sup>** | 15 (1.4) | 135 (1.3) | 4 (1.7) | 16 (0.7) |
| **Comorbidity score before MGUS/MBL diagnosis<sup>4</sup>** | **0** | 1262 (57.3) | 15975 (72.5) | 419 (74.7) | 4115 (73.4) |
|  | **1** | 510 (23.2) | 3630 (16.5) | 84 (15.0) | 898 (16.0) |
|  | **≥2** | 431 (19.6) | 2423 (11.0) | 58 (10.3) | 597 (10.6) |
|  |  | p<0.001 |  | p=0.78 |  |

<sup>1</sup>Followed-up until 15/03/2017; <sup>2</sup>Chronic lymphocytic leukaemia; <sup>3</sup>Prostate for men only, breast for women only; <sup>4</sup>≥1 hospital admission with a diagnosis of one of the 17 conditions included in the Charlson Comorbidity Index<sup>25-27</sup> during the 5 years to 1 month before MGUS/MBL diagnosis (patients)/pseudo-diagnosis (controls)

With respect to co-morbidity, in the years leading up to diagnosis MGUS patients were significantly more likely than their corresponding controls to have a record of at least one of the 17 co-morbidities specified in the Charlson Comorbidity Index[25–27] recorded in their discharge summaries; but no difference between MBL cases and their controls was evident (Table 1). More information about the extent of the differences between the hospital activity patterns of cases with MGUS/MBL and their general population controls, is shown in Figure 1; which shows inpatient and outpatient activity (excluding haematology) during the five-years before and the three-years after diagnosis of MGUS (Figure 1A) and MBL (Figure 1B). In the period before diagnosis, patients with MGUS (Figure 1A) had consistently higher outpatient activity rates than their controls, the disparity increasing markedly during the 18 months leading up to the formal diagnosis of MGUS by haematopathology. Although less pronounced, a similar pattern is evident in inpatient data. With smaller numbers and more scatter, variations in outpatient and inpatient activity in MBL are less evident (Figure 1B).

**Figure 1:**
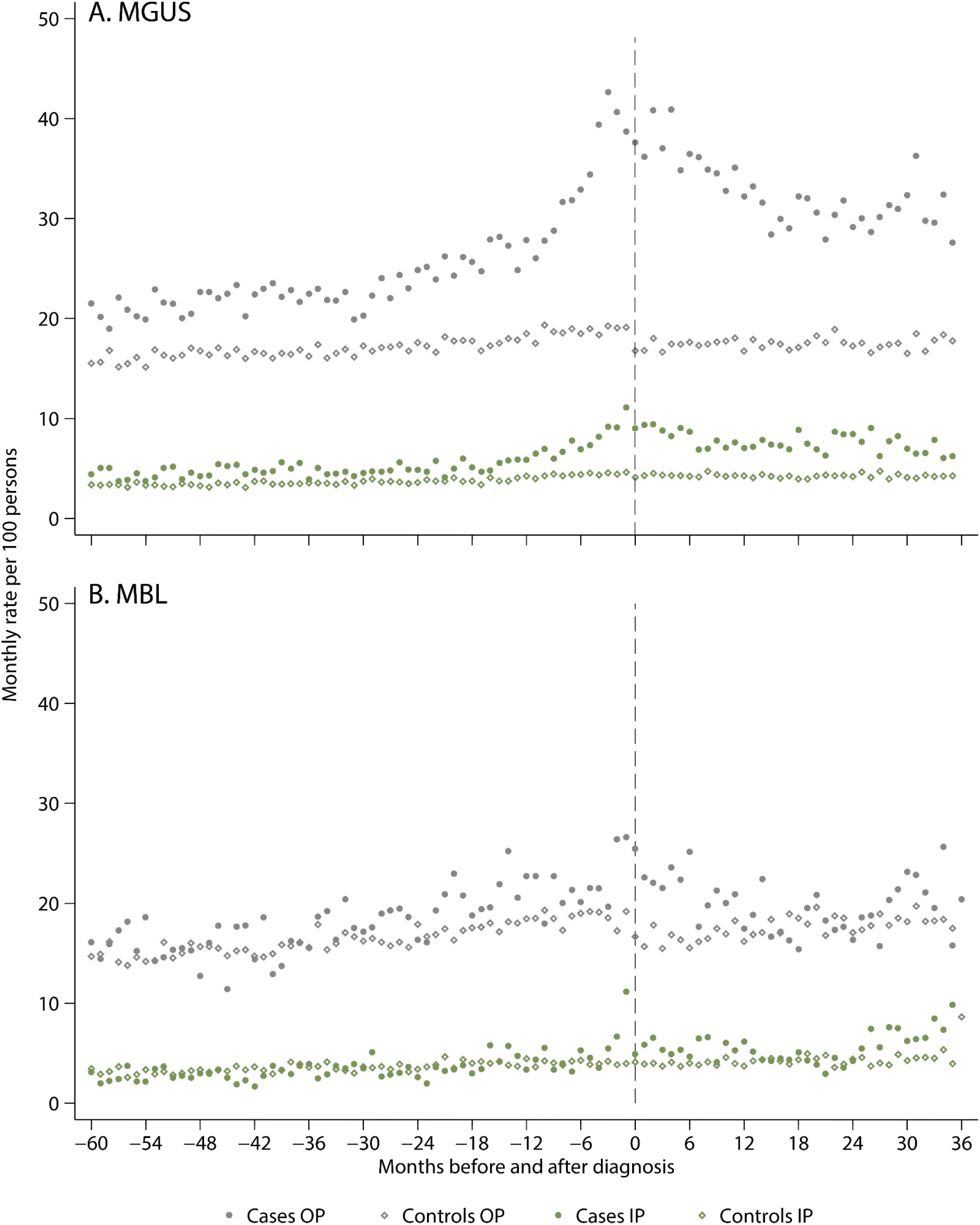
Inpatient (IP) and outpatient (OP) monthly hospital visit activity rates five years before to three years after diagnosis (cases)/pseudo-diagnosis (controls) excluding haematology attendances: A) monoclonal gammopathy of undetermined significance (MGUS) and B) monoclonal B-cell lymphocytosis (MBL) diagnosed 2009–15

Figure 2 shows outpatient attendance frequencies (at least two specialty-specific visits) in the three years before and in the three years after MGUS diagnosis for the top 25 clinical specialties; visits within one month (±) of diagnosis/pseudodiagnosis are excluded. As is evident from the plot, the increased outpatient activity seen among cases (Figure 1) occurs across a range of clinical specialties; the highest frequencies occurring in ophthalmology, haematology, general surgery, orthopaedics, general (internal) medicine and rheumatology. However, excluding haematology where, as expected, attendances increased markedly just before and after MGUS diagnosis, the largest rate ratios (RR) both before (Figure 3A) and after (Figures 3B) diagnosis were for nephrology (before diagnosis RR = 4.38, 95% CI 3.99–4.81; after diagnosis RR = 14.7, 95% CI 13.5–15.9) and rheumatology (before diagnosis RR = 3.38, 95% CI 3.16–3.61; after diagnosis RR = 5.45, 95% CI 5.09–5.83). Other significant associations (p<0.05) with rate ratio point estimates above 2.0 were evident for endocrinology, neurology, and respiratory medicine, as well as for the nurse-led monitoring activities which form part of on-going clinical care across a range of specialties.

**Figure 2:**
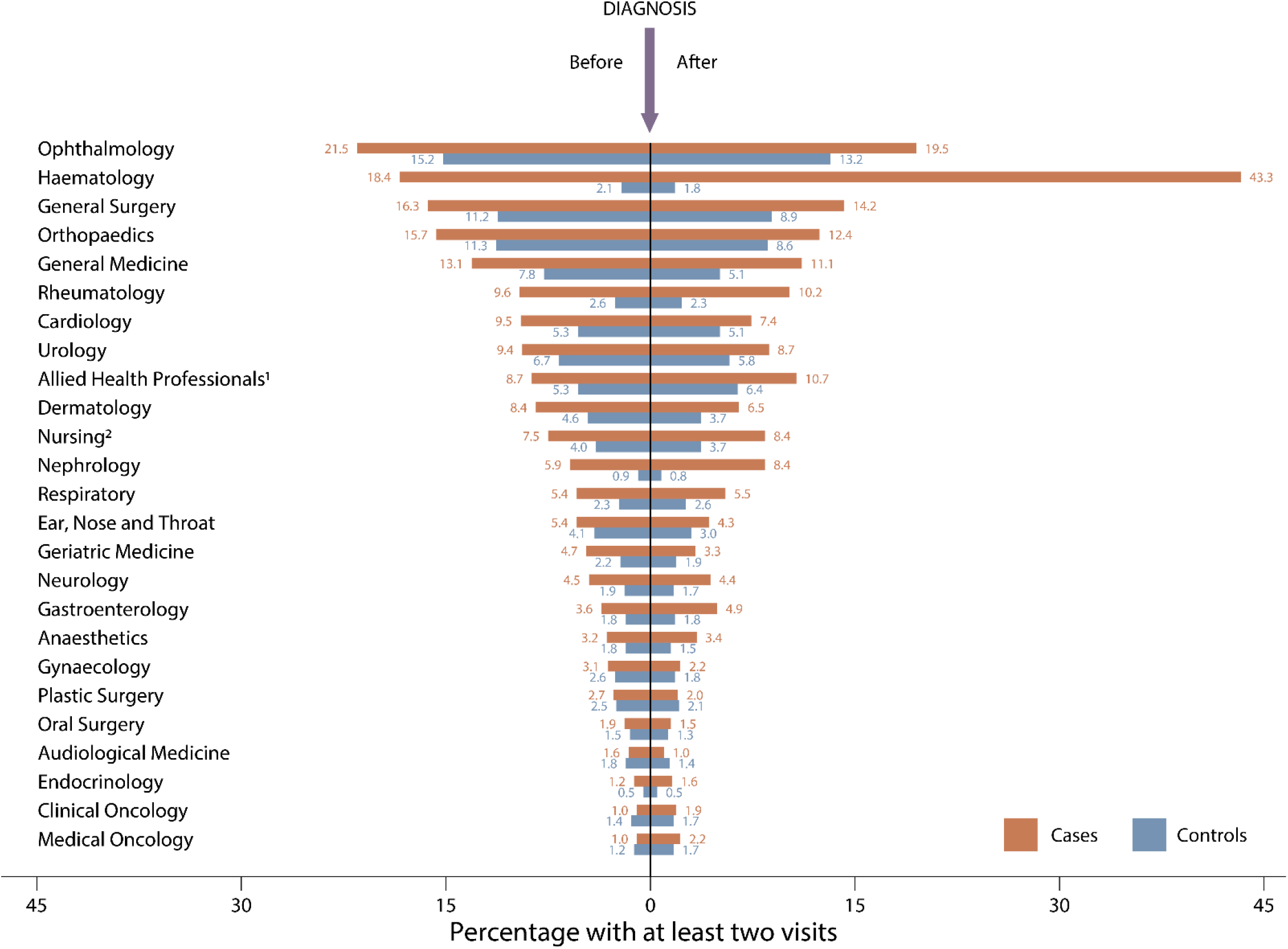
Percentage of cases and controls with at least two specialty-specific outpatient visits in the three years before and after diagnosis of monoclonal gammopathy of undetermined significance (MGUS). Top 25 recorded specialties, with visits within one month of diagnosis/pseudodiagnosis excluded.

**Figure 3:**
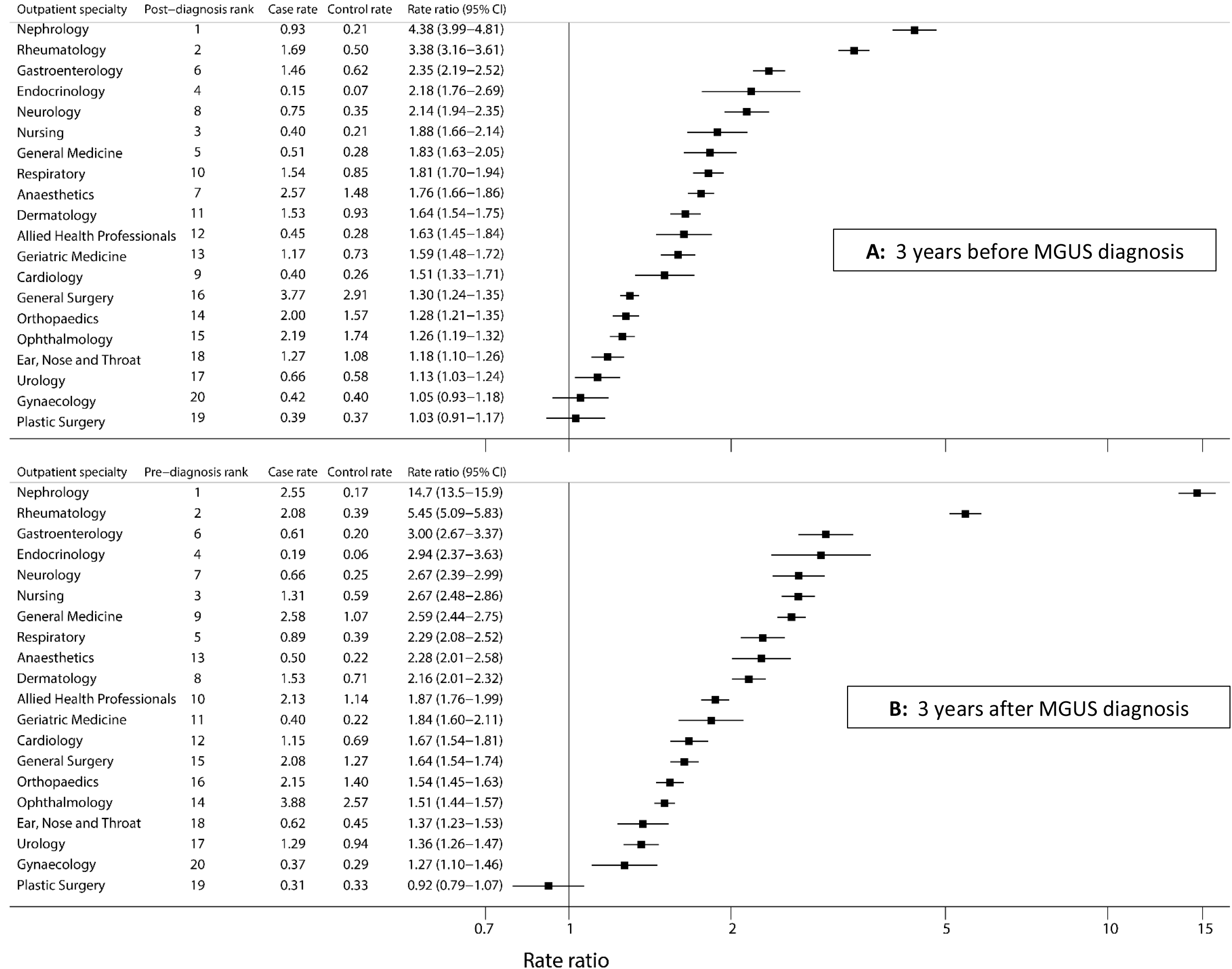
Monthly outpatient attendance rates (per 100 persons) in cases and controls and rate ratios by outpatient specialty with at least two visits (A) in the three years before diagnosis and (B) in the three years after diagnosis of monoclonal gammopathy of undetermined significance (MGUS)

MGUS data are stratified by subtype in Table 2. Accounting for around two-thirds of the total (n = 1469; 66.7%) the IgG subtype dominates, followed by IgM (n = 349; 15.8%) and IgA (n = 268; 12.2%). The remaining 117 (5.3%) ‘other’ category comprise a mix of subtypes: light chain only (n = 62), IgG + IgM (n = 17), IgG + IgA (n = 6), IgA + IgM (n = 1), IgE (n = 2), IgD (n = 1); and not recorded (n = 28). As expected, progression to myeloma in the three years following MGUS diagnosis was largely restricted to the IgG and IgA subtypes. The age distributions, five-year survival estimates (overall and relative) and non-haematological malignancy frequencies of the main subtypes were broadly similar; although patients in the combined ‘other’ category tended to be slightly older and to fare less well (RS = 75.1%, 95% CI 60.8–84.9%). The numbers of patients in the individual groups were, however, too sparse to examine the data in greater depth.

**Table 2:**
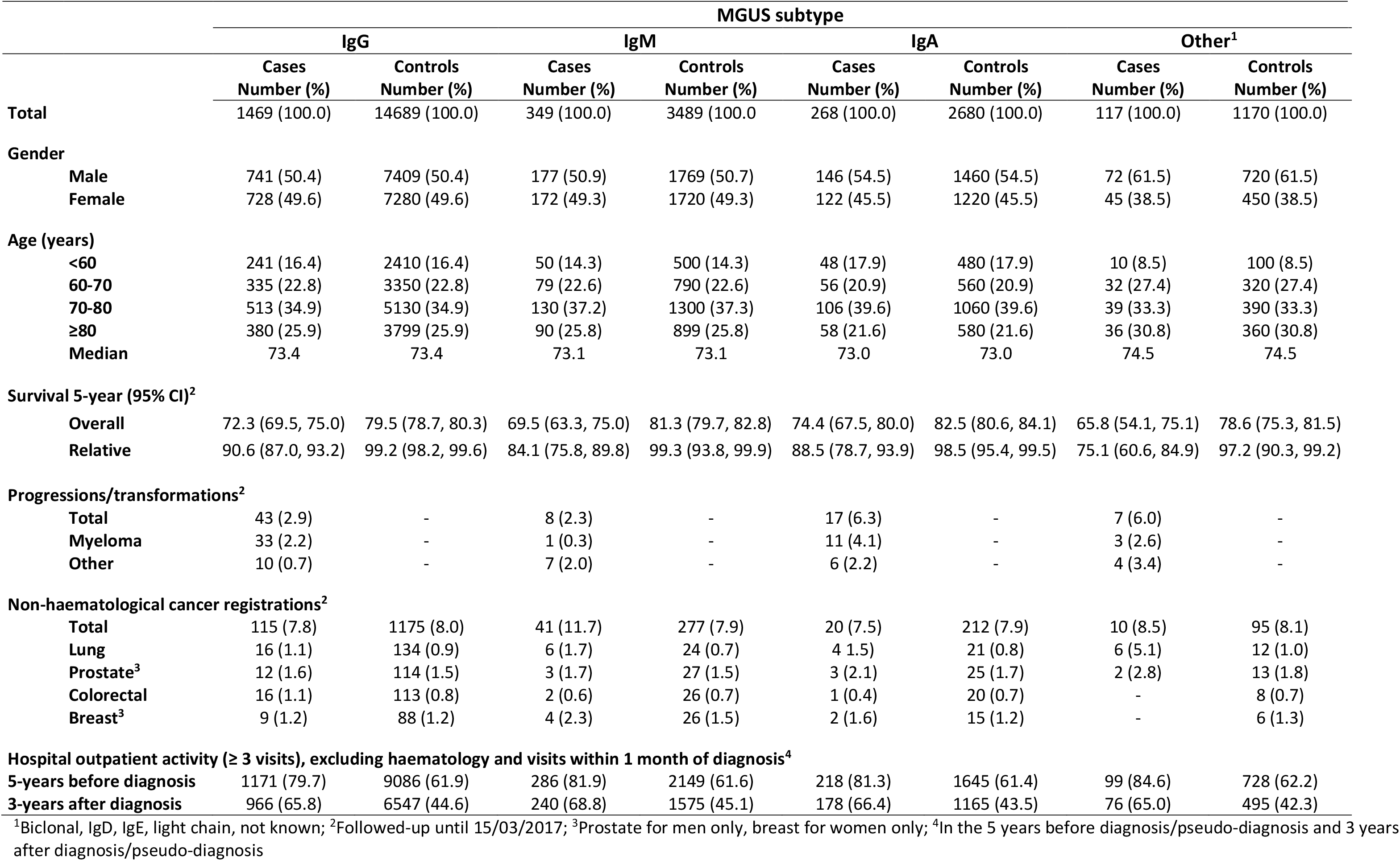
Characteristics of patients diagnosed with monoclonal gammopathy of undetermined significance (MGUS) and their corresponding controls, stratified by MGUS subtype; HMRN diagnoses 2009–2015

## DISCUSSION

Including data on nearly 3000 cases with pre-malignant clonal disorders and ten times as many age and sex-matched general population controls, this large UK record-linkage study found that individuals with MGUS not only experienced excess mortality and morbidity after diagnosis, but also excess morbidity in the 5 years before. By contrast, only marginal increases in mortality and morbidity were evident for MBL; none of which were consistent or varied significantly from the general population. Interestingly, progression patterns were in the opposite direction: in the years following detection of a pre-malignant clonal disorder, 3.4% (n = 75/2203) of those with MGUS developed a haematological malignancy (41 of which were myelomas) before April 2017, compared to 25.0% (n = 140/561) of those with MBL (137 of which were CLLs).

The elevated mortality and morbidity following a diagnosis of MGUS, which seems largely independent of progression to cancer, is consistent with reports relating to the potential clinical significance of this disorder.[11–18] In addition, our findings not only demonstrate the range and strength of the morbidity effects, but also show that levels are elevated many years before MGUS is diagnosed: excesses being observed in specialties covering most organ and tissue systems including nephrology, endocrinology, neurology, rheumatology, gastroenterology, dermatology and respiratory medicine. With respect to MBL, even though a quarter of patients developed CLL during the follow-up period, no effect on survival was detected. This result is, however, similar to that reported by other studies that used age and sex-matched controls.[3,30] Furthermore, although hospital activity increased in the months around the time of MBL diagnosis, no consistent differences or patterns before or after then, were detected. The number of patients in our MBL cohort (n = 561), although comparatively large is nonetheless smaller than the number of patients in our MGUS cohort (n = 2203). Furthermore, given the fact that MBL has been associated with increased susceptibility to infection and non-CLL related mortality[4,15,31], it is possible that the findings relating to subsequent morbidity and mortality could change as our data mature, length of follow-up increases, and linkage to primary care data occurs.

The age and sex distributions of our population-based cohorts are broadly similar to those of other published MBL[3,32,33] and MGUS[1,2,11] series; as is the dominance of the IgG MGUS subtype.[1,2] Providing nationally generalizable data, additional strengths of our study include its large well-defined population, within which all haematological malignancies and related clonal disorders are diagnosed, monitored, and coded using up-to-date standardized procedures at a central haematopathology laboratory.[19] In this context, it is important to bear in mind that most people with premalignant clonal disorders remain asymptomatic, and that our cohorts contain a relatively large proportion of people who came to clinical attention in primary and/or secondary care. Hence, our patient cohorts are likely to represent the more severe end of the disease spectrum; prevalence comparisons with population screening studies suggesting that this could be as low as 5% of the those ≥50 years with monoclonal immunoglobulin in their blood/urine.[5,20,33–35] Furthermore, during the study period (2009–15), in addition to the detection of a paraprotein in peripheral blood, around 70% (1546/2203) of MGUS patients in our cohort had a confirmatory bone marrow sample. The patient characteristics and secondary care activity patterns of those who had a bone marrow sample were, however, broadly similar to those who did not (data not shown).

The diversity of morbidity effects seen among individuals with MGUS is consistent with the expanding body of evidence relating to the potential adverse impact that even low levels of circulating monoclonal protein (M-protein) can have. Thus far, the complex underpinning mechanisms identified include; deposition of M-protein aggregations of varying immunoglobulin subtypes in different organs, as well as the induction of auto-antibodies and cytokines that can impact on organs and tissues in a variety of deleterious ways.[13,18,36,37] Indeed, the recognized number of M-protein mediated entities is increasing, with several affecting multiple organs; well-known deposition syndromes including primary amyloidosis and paraneoplastic conditions such as POEMS syndrome (polyneuropathy, organomegaly, endocrinopathy, monoclonal gammopathy, and skin changes).[7,38] As evidenced in our analysis, kidney involvement is frequent, both in the years before (four-fold excess) and after (fifteen-fold excess) MGUS diagnosis. Indeed, the umbrella term monoclonal gammopathy of renal significance (MGRS) has recently been suggested to cover all M-protein mediated kidney disorders that fail to meet the diagnostic criteria for multiple myeloma or any other B-cell malignancy.[13,18,39] Other organ-specific terms continue to emerge, and with a view to improving recognition of these complex disorders which clearly pose significant diagnostic and treatment challenges, the overarching term monoclonal gammopathy of clinical significance (MGCS) has also been suggested.[18]

From a haematological malignancy perspective, MGUS and MBL are generally considered to be relatively benign conditions. However, both can have other deleterious health consequences; the effect of monoclonal gammopathy being particularly striking. Impacting significantly on survival and having the potential to cause systemic disease and wide-ranging damage to most organs and tissues, the adverse outcomes associated with the M-proteins produced by the abnormal B-cell clone can be severe and extend over many years. Even though most people with monoclonal immunoglobulins never develop a B-cell malignancy or suffer from any other form of M-protein related organ/tissue related disorder, the consequences for those that do can be extremely serious. In this regard, early targeting of pathogenic B-cell clones could mitigate both cancer and non-cancer effects; but currently, although knowledge is increasing, there is no known way to reliably identify such clones in the absence of other signs/symptoms. Hence, population screening cannot be recommended, and diagnosis remains reliant on clinical suspicion. However, the long-standing nature of the co-morbidity associations seen prior to MGUS diagnosis in our data, suggest that there may be room for improvement. Hence, in the future, the implementation of strategies to improve awareness and earlier detection, as well as monitoring of high-risk patient groups, could prove beneficial.

## Data Availability

Ethical approvals and data restrictions mean that data cannot be shared; but collaborative projects can be undertaken. The corresponding author can be contacted for more information.

## ARTICLE SUMMARY

### Strengths and limitations of this study

- Data are from an established population-based cohort within which all haematological malignancies and related clonal disorders are diagnosed, monitored, and coded using up-to-date procedures at a central haematopathology laboratory.
- Providing nationally generalizable data, all diagnoses are included, and complete follow-up is achieved via linkage to nationwide information on mortality and morbidity.
- The age and sex-matched general population cohort enables baseline activity and rate ratios to be calculated, both before and after premalignancy detection.
- Most people with premalignant clonal disorders are asymptomatic, and our patient cohort contains a relatively large proportion of people who came to clinical attention.
- Analyses are constrained by the fact that data that hospital episodes statistics are primarily collected for administrative and clinical purposes, and not for research.

### Funding

This work was supported by Cancer Research UK, grant numbers 18362 and 29685; and Blood Cancer UK, grant number 15037

### Competing interests

None

## Ethics Approval

The Haematological Malignancy Research Network has ethics approval (REC 04/01205/69) from Leeds West Ethics Committee, R&D approval from each NHS Trust, and exemption from Section 251 of the Health & Social Care Act (PIAG 1–05 9h)/2007)

## Contributors

ER, AS, DH and RP initiated the patient cohort within which this research is nested; and ER, AS, and EK designed the comparison cohort. ER, AS and ML planned this study and analyses. R de T and AR oversaw all laboratory procedures; and ML, DP, EK, TB managed the data and carried out data analysis. GC, RP, AR, and RN commented on the biological and clinical aspects. ER and ML drafted the manuscript, which was approved by all authors

---

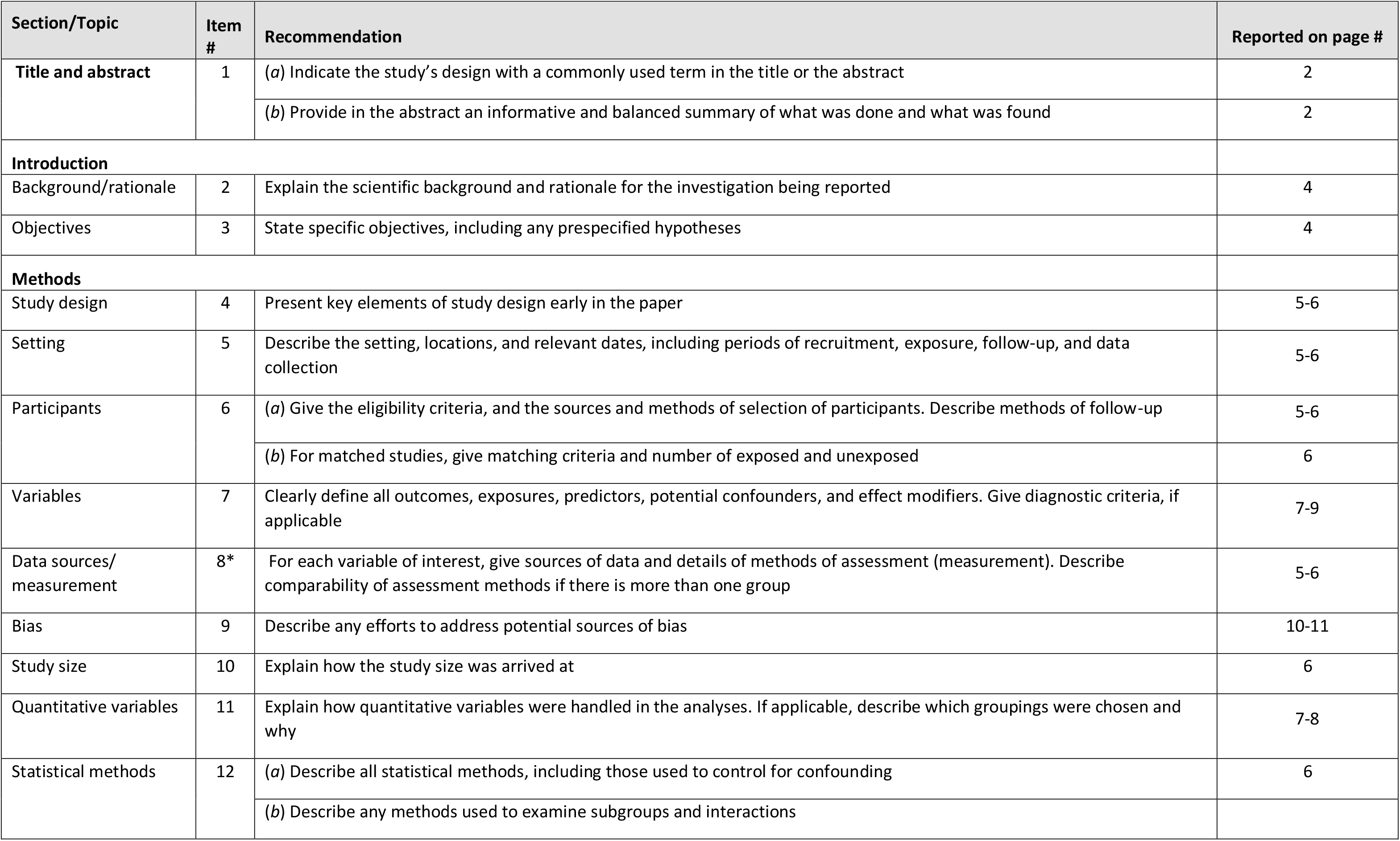
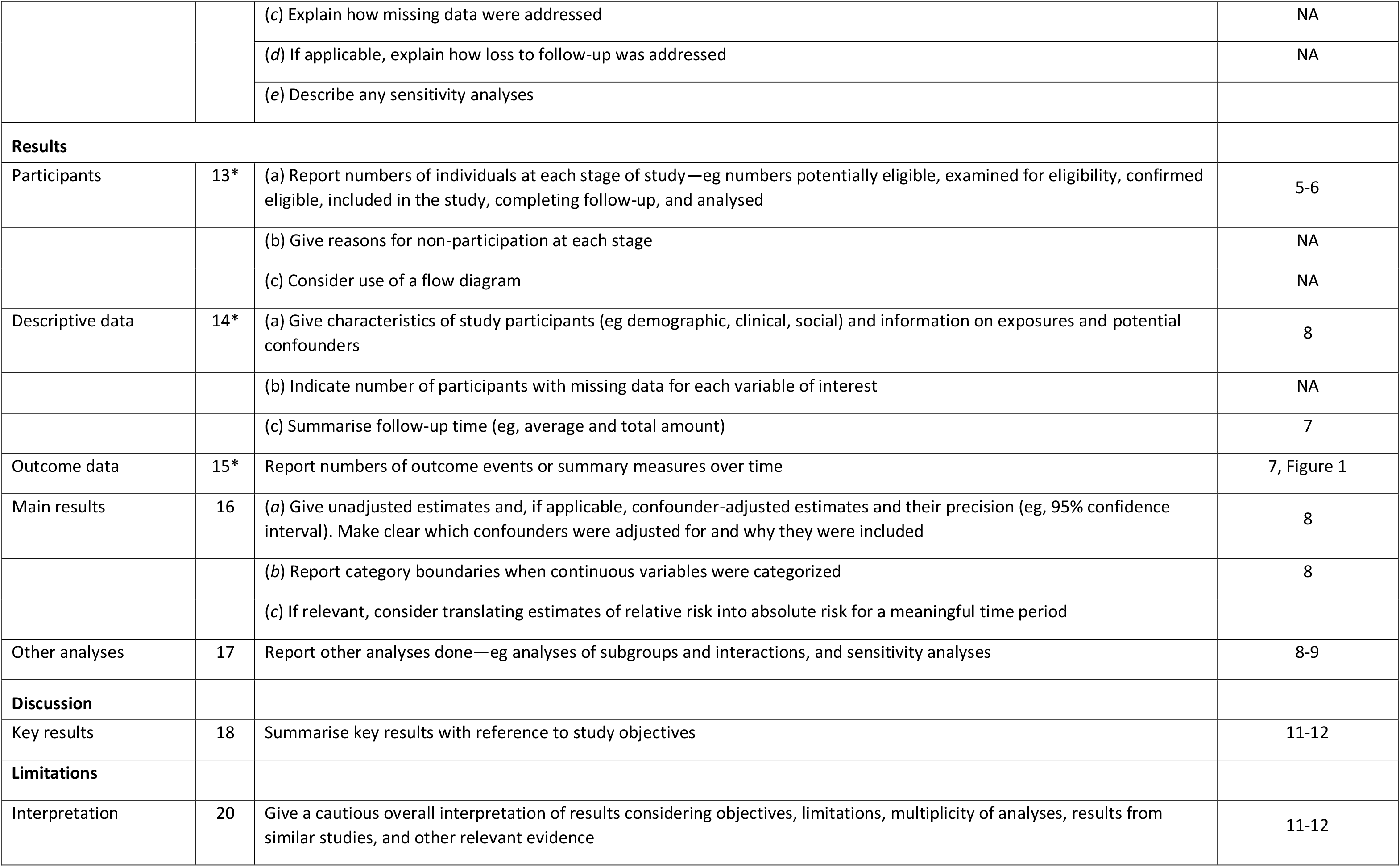
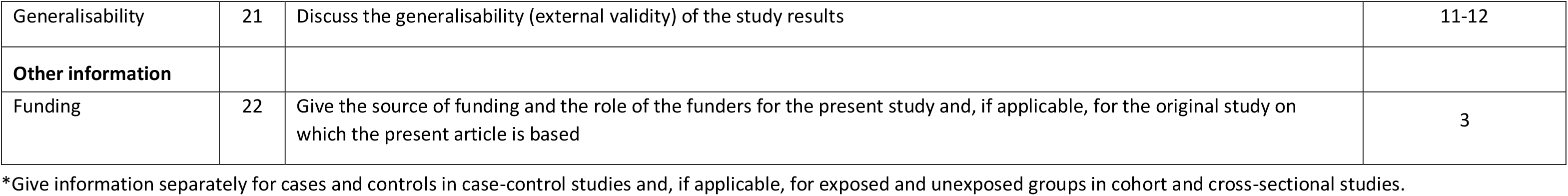
STROBE 2007 (v4) Statement—Checklist of items that should be included in reports of *cohort studies*

| Section/Topic | Item # | Recommendation | Reported on page # |
| --- | --- | --- | --- |
| Title and abstract | 1 | (a) Indicate the study’s design with a commonly used term in the title or the abstract | 2 |
|  |  | (b) Provide in the abstract an informative and balanced summary of what was done and what was found | 2 |
| Introduction |  |  |  |
| Background/rationale | 2 | Explain the scientific background and rationale for the investigation being reported | 4 |
| Objectives | 3 | State specific objectives, including any prespecified hypotheses | 4 |
| Methods |  |  |  |
| Study design | 4 | Present key elements of study design early in the paper | 5-6 |
| Setting | 5 | Describe the setting, locations, and relevant dates, including periods of recruitment, exposure, follow-up, and data collection | 5-6 |
| Participants | 6 | (a) Give the eligibility criteria, and the sources and methods of selection of participants. Describe methods of follow-up | 5-6 |
|  |  | (b) For matched studies, give matching criteria and number of exposed and unexposed | 6 |
| Variables | 7 | Clearly define all outcomes, exposures, predictors, potential confounders, and effect modifiers. Give diagnostic criteria, if applicable | 7-9 |
| Data sources/<br>measurement | 8* | For each variable of interest, give sources of data and details of methods of assessment (measurement). Describe comparability of assessment methods if there is more than one group | 5-6 |
| Bias | 9 | Describe any efforts to address potential sources of bias | 10-11 |
| Study size | 10 | Explain how the study size was arrived at | 6 |
| Quantitative variables | 11 | Explain how quantitative variables were handled in the analyses. If applicable, describe which groupings were chosen and why | 7-8 |
| Statistical methods | 12 | (a) Describe all statistical methods, including those used to control for confounding | 6 |
|  |  | (b) Describe any methods used to examine subgroups and interactions |  |
|  |  | (c) Explain how missing data were addressed | NA |
|  |  | (d) If applicable, explain how loss to follow-up was addressed | NA |
|  |  | (e) Describe any sensitivity analyses |  |
| **Results** |  |  |  |
| Participants | 13* | (a) Report numbers of individuals at each stage of study—eg numbers potentially eligible, examined for eligibility, confirmed eligible, included in the study, completing follow-up, and analysed | 5-6 |
|  |  | (b) Give reasons for non-participation at each stage | NA |
|  |  | (c) Consider use of a flow diagram | NA |
| Descriptive data | 14* | (a) Give characteristics of study participants (eg demographic, clinical, social) and information on exposures and potential confounders | 8 |
|  |  | (b) Indicate number of participants with missing data for each variable of interest | NA |
|  |  | (c) Summarise follow-up time (eg, average and total amount) | 7 |
| Outcome data | 15* | Report numbers of outcome events or summary measures over time | 7, Figure 1 |
| Main results | 16 | (a) Give unadjusted estimates and, if applicable, confounder-adjusted estimates and their precision (eg, 95% confidence interval). Make clear which confounders were adjusted for and why they were included | 8 |
|  |  | (b) Report category boundaries when continuous variables were categorized | 8 |
|  |  | (c) If relevant, consider translating estimates of relative risk into absolute risk for a meaningful time period |  |
| Other analyses | 17 | Report other analyses done—eg analyses of subgroups and interactions, and sensitivity analyses | 8-9 |
| **Discussion** |  |  |  |
| Key results | 18 | Summarise key results with reference to study objectives | 11-12 |
| **Limitations** |  |  |  |
| Interpretation | 20 | Give a cautious overall interpretation of results considering objectives, limitations, multiplicity of analyses, results from similar studies, and other relevant evidence | 11-12 |
| Generalisability | 21 | Discuss the generalisability (external validity) of the study results | 11-12 |
| **Other information** |  |  |  |
| Funding | 22 | Give the source of funding and the role of the funders for the present study and, if applicable, for the original study on which the present article is based | 3 |

\*Give information separately for cases and controls in case-control studies and, if applicable, for exposed and unexposed groups in cohort and cross-sectional studies.

## Notes

### Competing Interest Statement

The authors have declared no competing interest.

### Author Declarations

The Haematological Malignancy Research Network has ethics approval (REC 04/01205/69) from Leeds West Ethics Committee, R&D approval from each NHS Trust, and exemption from Section 251 of the Health & Social Care Act (PIAG 1-05 9h)/2007)

